# Risk factors and cognitive correlates of white matter hyperintensities in ethnically diverse populations without dementia: the COSMIC consortium

**DOI:** 10.1101/2023.08.30.23294876

**Authors:** Keshuo Lin, Wei Wen, Darren M. Lipnicki, Louise Mewton, Rory Chen, Jing Du, Dadong Wang, Ingmar Skoog, Therese Rydberg Sterner, Jenna Najar, Ki Woong Kim, Ji Won Han, Jun Sung Kim, Tze Pin Ng, Roger Ho, Denise Qian Ling Chua, Kaarin J. Anstey, Nicolas Cherbuin, Moyra E. Mortby, Henry Brodaty, Nicole Kochan, Perminder S. Sachdev, Jiyang Jiang, Cohort Studies of Memory in an International Consortium (COSMIC)

**Author notes:** **Corresponding author:** Dr. Jiyang Jiang. Level 1, AGSM building (G27), UNSW Kensington, Sydney NSW 2033, Australia. E.

## Abstract

**INTRODUCTION:** White matter hyperintensities (WMH) are an important imaging marker for cerebral small vessel diseases, but their risk factors and cognitive associations have not been well-documented in populations of different ethnicities and/or from different geographical regions.

**METHOD:** Magnetic resonance imaging data of five population-based cohorts of non-demented older individuals from Australia, Singapore, South Korea, and Sweden (N = 1,946) were examined for WMH and their associations with vascular risk factors and cognition.

**RESULT:** Factors associated with larger whole brain WMH volumes included diabetes, hypertension, stroke, current smoking, body mass index, higher alcohol intake and insufficient physical activity. Participants with moderate or higher physical activity had less WMH than those who never exercised, but the former two groups did not differ. Hypertension and stroke had stronger associations with WMH volumes in the White, compared to Asian subsample.

**DISCUSSION:** The current study highlighted the ethnic differences in the contributions of vascular risk factors to WMH.

## Introduction

White matter hyperintensities (WMH) are an important imaging marker for cerebrovascular diseases. A rich body of research has examined risk factors for WMH (1, 2) and associations between WMH and cognition (3, 4). In particular, studies have found a heavier burden of WMH with hypertension (5), higher blood pressure in normal range (6, 7), diabetes (8), cardiovascular disease (9), excessive alcohol consumption (10), and insufficient physical activity (3, 10, 11). Poorer performance in the cognitive domains of processing speed (1) and executive function (12) has also been associated with WMH. Further, a meta-analysis of 22 longitudinal studies found that WMH accumulations were associated with increased dementia and stroke risks, and mortality (3).

Ethnicity differences in the prevalence of WMH, and their associations with risk factors and cognition, have not been well documented, although studies in different ethnicities exist. For example, a population-based study of 688 elderly Japanese reported that higher systolic blood pressure, current drinking, lower total cholesterol, and poorer cognitive function were associated with higher WMH volumes (13). A study of 1,748 population-based whites showed that higher WMH volumes were associated with more cigarette smoking per day, a higher prevalence of cardiovascular disease, poorer memory, and reduced organization skills (14, 15). The current study aimed to examine associations of WMH with risk factors and cognition, and ethnicity differences in their associations, by pooling magnetic resonance imaging (MRI) data from 5 population-based studies of non-demented older adults in Cohort Studies of Memory in an International Consortium (COSMIC; (16)).

The primary advantage of using COSMIC data is the availability of population-based, community-dwelling older individuals from different countries in the world, with participant-level clinical and MRI data available for analyses. The major challenge arises from the need to harmonize data pertaining to risk factors, neuropsychological assessments, and MRI. To address these challenges, COSMIC has introduced comprehensive protocols, and the harmonization of risk factors and neuropsychological data has been largely achieved (17).

## Aims

To examine the associations of WMH volumes with vascular risk factors and cognition, and ethnicity differences in their associations, in a pooled sample of population-based studies of non-demented individuals from four countries.

## Methods

### Participating studies

Five COSMIC studies with MRI scans were included in the current study, viz., Korean Longitudinal Study on Cognitive Aging and Dementia (KLOSCAD) (18), the Gothenburg H70 Birth Cohort Study (H70-study) (19), Singapore Longitudinal Aging Studies (SLAS-I and SLAS-II) (20), Sydney Memory and Ageing Study (MAS) (21), and The Personality and Total Health (PATH) Through Life study (60 years and above) (22). Baseline data were used for all studies except PATH to maximise the sample size. Wave 2 data of PATH were included because cognitive data to assess executive function and processing speed were not available at baseline. Five participants from KLOSCAD and 27 from SLAS with dementia were excluded.

### MRI acquisition

We analysed T1- and T2-weighted Fluid Attenuated Inversion Recovery (FLAIR) scans acquired across 7 MRI scanners. KLOSCAD and H70-study used Philips 3T Achieva scanners (Philips Medical Systems, The Netherlands) (19, 23), and PATH used a Philips 1.5T Gyroscan scanner (22). Of the 474 MAS participants, 240 were scanned with a Philips 3T Intera Quasar scanner, and the remaining 234 with a Philips 3T Achieva Quasar Dual scanner (21). SLAS-I and SLAS-II used a Siemens 3T Tim Trio and a GE Healthcare 1.5T HDXT scanner, respectively. Detailed imaging acquisition parameters are summarised in Supplementary Table 1.

### WMH segmentation and quantification

WMH volumes were quantified using UBO detector (24). Briefly, FLAIR scans were linearly registered to T1 images. Tissue segmentation was then conducted, and the warp field from native T1 space to Diffeomorphic Anatomical Registration Through Exponentiated Lie Algebra (DARTEL; (25)) space was generated. The warp fields were applied to FLAIR images in native T1 spaces to bring FLAIR scans to the DARTEL space. After removing non-brain tissue, FSL FAST was applied to segment DARTEL-space FLAIR into candidate clusters. By considering intensity, anatomical location, and cluster size features, a *k*-nearest neighbour classifier was applied to separate non-WMH from WMH clusters. WMH voxels <12 mm from lateral ventricles were considered as periventricular WMH (PVWMH) voxels, and those ≥12 mm were classified as deep WMH (DWMH) voxels. Quality control was conducted by visualising the resultant WMH masks superimposed onto corresponding FLAIR images. Seventy-eight scans were excluded due to inaccurate segmentations (Supplementary Table 2).

### Harmonisation of non-imaging variables

The harmonisation of body mass index (BMI), diabetes, hypertension, hypercholesterolaemia, atrial fibrillation (AF), cardiovascular disease, stroke, physical activity, and smoking, followed the protocols in a previous COSMIC study ((17); Supplementary Tables 3-7). Physical activity was categorised as inactive (scored as 0), occasional exercise (scored as 1) and frequent (scored as 2) (Supplementary Tables 3-7).

Since SLAS only acquired data for the frequency of alcohol drinking, and 97.1% of participants reported never/rarely drinking, it was excluded from the analyses of alcohol consumption. The harmonisation of alcohol consumption in other cohorts followed another COSMIC study ((26); Supplementary Tables 3-7). AF data were not acquired for PATH participants.

The harmonisation of global cognition (Mini-Mental State Examination (MMSE)), memory (delayed world list recall), language (semantic fluency - animals), processing speed (Trail Making Test A), and executive function (Trail Making Test B), also followed COSMIC protocols ((17); Supplementary Text 1). Trail Marking Test A and B completion times were multiplied by minus one for better interpretation. Processing speed and executive function data were not available for H70-study. For some SLAS participants, memory (N = 100), language (N = 115), processing speed (N = 120), and executive function (N = 138) data were not available.

### Harmonisation of WMH volumes

The comparison of different harmonisation methods for WMH volumes was summarised in Supplementary Text 2. Age effects and Kolmogorov-Smirnov tests were used to assess the performance of harmonisation. ComBat-harmonised WMH volumes were used for further analyses.

### Statistical analyses

Controlling for age and sex, the association between risk factors and WMH was examined with structural equation models (SEM) implemented in the *Lavaan* (27) package in R. Maximum likelihood estimator with robust standard errors and a scaled test statistic were used. Full Information Maximum Likelihood was used to deal with missing values. To test the associations of risk factors on WMH, all vascular risk factors were used as independent variables of interest. Differences in the effect of each risk factor on PVWMH and DWMH were also examined with SEM. The associations between cognition and WMH volumes, and the mediation effects of WMH volumes in the associations between vascular risk factors and cognition, were also examined in SEM (details see Supplementary Text 3). The moderating effects of ethnicity (3 White and 2 Asian cohorts) in the association between vascular risk factors and WMH were tested. For associations where the interaction term between ethnicity and risk factor had a p-value less than 0.1, we ran regression analyses in White and Asian subsamples separately to investigate the ethnicity differences. In the tests for ethnicity differences, only White and Asian participants were included. Non-White (21 MAS and 12 PATH participants) and non-Asian (1 SLAS participant) individuals were excluded. Outliers for WMH volumes and the cognitive scores were removed through applying a 3 standard deviation threshold.

## Results

### Sample characteristics

Participants were aged 56 to 94 years at the time of data acquisition, and 54.8% were female. The excluded participants (WMH quality control and WMH outliers, n = 125) were aged 62 to 74 (mean age is 71), and 55.2% were female. There were no differences in sex distribution between the excluded participants and the whole sample (χ^2^ = 0.018, p = 0.991). Sample characteristics and the details of each cohort exclusion are summarised in Table 1.

**Table 1.** Sample characteristics.

| Parameters |  | H70-study | KLOSCAD | MAS | PATH | SLAS | Total |
| --- | --- | --- | --- | --- | --- | --- | --- |
| **N (Total/QC/Outliers)<sup>a</sup>** |  | 704/700/668 | 260/257/255 | 497/474/468 | 414/384/380 | 196/178/175 | 2071/1994/1946 |
| **Age (mean [range])** |  | 71 [70,72] | 72 [59,89] | 79 [70,90] | 67 [64,70] | 74 [56,94] | 72 [56, 94] |
| **Sex (female)** |  | 52.9% | 57.6% | 54.2% | 43.8% | 74.8% | 54.8% |
| **Diabetes** |  | 15.7% | 43.1% | 12.9% | 9.4% | 21.3% | 18.1% |
| **Hypertension** |  | 74.2% | 85.5% | 80.4% | 64.3% | 69.0% | 74.6% |
| **Hypercholesterolemia** |  | 47.1% | 74.0% | 67.7% | 36.3% | 61.6% | 55.2% |
| **Atrial fibrillation** |  | 4.1% | 6.1% | 4.3% | - | 3.5% | 4.4% |
| **Cardiovascular disease** |  | 6.9% | 19.1% | 22.8% | 14.1% | 10.5% | 13.9% |
| **Stroke** |  | 8.3% | 8.0% | 2.3% | 0.0% | 29.1% | 8.0% |
| **Current smoking** |  | 7.7% | 5.3% | 3.4% | 6.8% | 28.7% | 8.9% |
| **Physical activity (occasional)** |  | 11.9% | 66.4% | 50.1% | 72.1% | 81.1% | 45.3% |
| **Physical activity (frequent)** |  | 85.7% | 12.2% | 34.2% | 18.3% | 7.6% | 44.3% |
| **Alcohol (drinks/week)<sup>b</sup>** |  | 8 [0, 68.7] | 2 [0, 42] | 6.3 [0, 56] | 3.8 [0, 63.4] | - | 3.8 [0, 68.7] |
| **BMI** |  | 26.0±4.4 | 23.5±2.8 | 26.7±4.2 | 26.6±4.2 | 23.4±4.1 | 26.0±4.3 |
| **Education (years)** |  | 13.3±4.1 | 12.1±4.5 | 11.7±3.6 | 14.0±2.8 | 3.6±3.7 | 12.0±4.7 |
| **Cognition** | **MMSE** | 29±1.3 | 26.8±3 | 28.9±1.4 | 29.1±1.4 | 25.1±4.4 | 28.3±2.5 |
|  | **Memory** | 8.6±1.6 | 5.4±2.4 | 7.5±3.5 | 6.0±2.5 | 5.4±3.4 | 6.8±3.1 |
|  | **Language** | 24.2±6.1 | 16.7±5.1 | 15.8±4.6 | - | 12.1±2.8 | 19.5±4.7 |
|  | **Processing Speed** | - | 57.6±46 | 46±16.1 | 34.4±11.0 | 103.7±51.3 | 50.5±34.9 |
|  | **Executive**<br>**Function** | - | 163.8±87 | 117.9±55.6 | 79.8±30.4 | 184.4±61.5 | 121.5±68.6 |

### Associations between vascular risk factors and WMH

The associations between vascular risk factors and WMH volumes are summarised in Table 2. Hypertension (β = 0.056 - 0.087, p = 0.001 – 0.005), current smoking (but not past smoking; β = 0.096 – 0.114, p = 0.001 – 0.007) and higher BMI (β = 0.005 – 0.006, p = 0.006 – 0.009) were associated with higher whole brain WMH (WBWMH), PVWMH and DWMH volumes. Higher number of drinks per week (β = 0.005, p = 0.031 – 0.037) and insufficient physical activity (β = 0.028 – 0.032, p = 0.020 – 0.039) were associated with WBWMH and PVWMH volumes. Participants with diabetes had higher WBWMH volumes (β = 0.043, p = 0.047), especially in the periventricular regions (β = 0.051, p = 0.022). History of stroke was associated with higher WBWMH volumes (β = 0.085, p = 0.039), especially in deep white matter regions (β = 0.143, p = 0.009). The associations between stroke and DWMH volumes were significantly stronger than the associations between stroke and PVWMH volumes (p = 0.004).

**Table 2.** Associations between vascular risk factors and harmonised WMH volumes.

|  | Whole Brain WMH |  |  | Periventricular WMH |  |  | Deep WMH |  |  |
| --- | --- | --- | --- | --- | --- | --- | --- | --- | --- |
| | $\beta$ | CI lower | CI upper | $\beta$ | CI lower | CI upper | $\beta$ | CI lower | CI upper |
| Diabetes | 0.043* | 0.001 | 0.087 | 0.051* | 0.007 | 0.095 | 0.037 | -0.025 | 0.098 |
| hypertension | 0.059** | 0.021 | 0.098 | 0.056** | 0.005 | 0.096 | 0.087*** | 0.034 | 0.140 |
| Hypercholesterolemia | -0.025 | -0.062 | 0.013 | -0.025 | -0.063 | 0.013 | -0.022 | -0.075 | 0.031 |
| Atrial fibrillation | 0.012 | -0.083 | 0.107 | 0.010 | -0.084 | 0.104 | -0.013 | -0.160 | 0.135 |
| cardiovascular disease | 0.033 | -0.015 | 0.082 | 0.021 | -0.028 | 0.070 | 0.051 | -0.016 | 0.144 |
| Stroke | 0.085* | 0.004 | 0.146 | 0.043 | -0.007 | 0.107 | 0.143** | 0.033 | 0.254 |
| Smoking | 0.096** | 0.035 | 0.157 | 0.099*** | 0.039 | 0.159 | 0.114** | 0.031 | 0.198 |
| Body Mass Index | 0.005** | 0.001 | 0.011 | 0.006** | 0.001 | 0.012 | 0.005** | 0.001 | 0.008 |
| Physical activity | -0.032* | -0.059 | -0.005 | -0.028* | -0.055 | -0.001 | -0.035 | -0.074 | 0.003 |
| Alcohol consumption | 0.005* | <0.001 | 0.009 | 0.005* | <0.001 | 0.009 | 0.003 | -0.001 | 0.012 |

The table lists all the regression coefficients for the risk factors. \* - p-value <0.05, \*\* - p-value <0.01, \*\*\* - p-value < 0.001. CI - 95% confidence interval

The differences in WMH volumes between different levels in smoking and physical activity are summarized in Figure 1. As shown in Figure 1, the frequent and occasional exercise groups had less WBWMH (mean difference = 0.092 - 0.106, p = 0.002 – 0.016) and DWMH (mean difference = 0.083 - 0.093, p = 0.037 – 0.045) volumes than the inactive group. No significant differences were found between the frequent and occasional exercise groups (p > 0.805).

**Figure 1.**
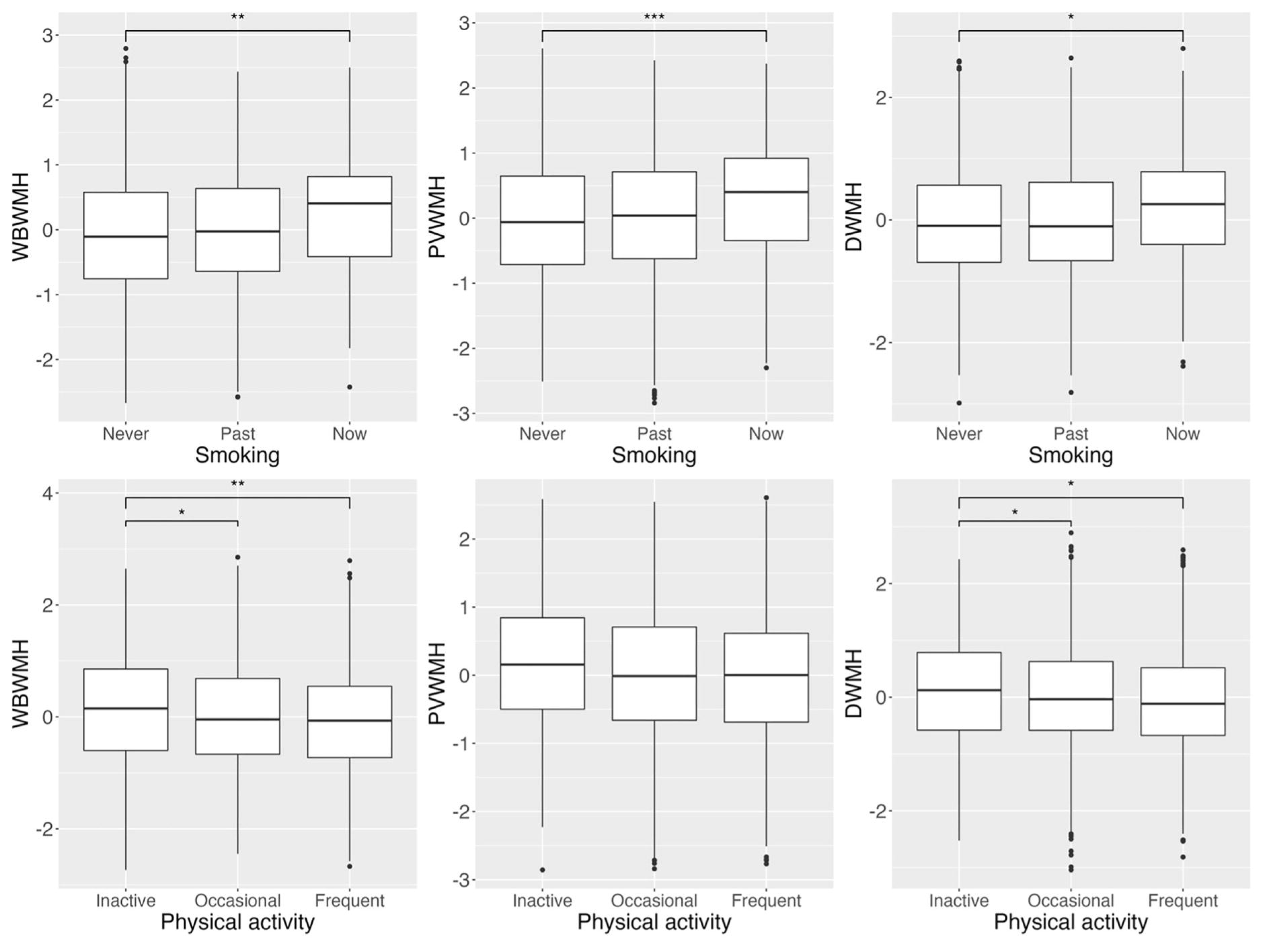
– The differences in WMH volumes between different levels of physical activities (first row) and smoking (second row). * - p-value <0.05, **-p-value <0.01, ***-p-value < 0.001. WBWMH – whole brain white matter hyperintensities; PVWMH – periventricular white matter hyperintensities; DWMH – deep white matter hyperintensities.

### Associations between WMH and cognition, and mediation effects of WMH

Associations between WMH volumes and cognition are summarised in Table 3. Higher DWMH volumes were associated with worse performance in the processing speed domain (β = −0.067, p = 0.026). Higher WBWMH (β = −0.057, p = 0.040) and DWMH (β = −0.063, p = 0.037) volumes were associated with worse performance in the executive function domain. No mediation effects of WMH were found in the associations between vascular risk factors and cognition (Supplementary Table 8).

**Table 3.** Associations between WMH volumes and cognition.

|  | Whole Brain WMH |  |  | Periventricular WMH |  |  | Deep WMH |  |  |
| --- | --- | --- | --- | --- | --- | --- | --- | --- | --- |
| **Cognition** | $\beta$ | CI lower | CI upper | $\beta$ | CI lower | CI upper | $\beta$ | CI lower | CI upper |
| **Mini-Mental State Examination** | -0.040 | -0.082 | 0.002 | -0.027 | -0.069 | 0.014 | -0.033 | -0.102 | 0.037 |
| **Memory** | -0.035 | -0.105 | 0.034 | -0.040 | -0.110 | 0.030 | -0.039 | -0.108 | 0.029 |
| **Language** | -0.020 | -0.088 | 0.046 | -0.015 | -0.083 | 0.052 | -0.024 | -0.091 | 0.041 |
| **Processing Speed** | -0.049 | -0.107 | 0.008 | -0.043 | -0.099 | 0.013 | -0.067* | -0.137 | -0.003 |
| **Executive Function** | -0.057* | -0.113 | <0.001 | -0.041 | -0.098 | 0.015 | -0.063* | -0.127 | <0.001 |

The table lists all the regression coefficients for cognitive variables. P-values for the associations were shown as \* - p-value <0.05. CI - 95% confidence interval.

### Moderation effects of ethnicity in the associations between vascular risk factors and WMH

The differences in WMH volumes between White and Asian groups are summarized in Figure 2, but were not statistically significant when adjusting for age and sex (mean differences < 0.002, p-value > 0.305).

**Figure 2.**
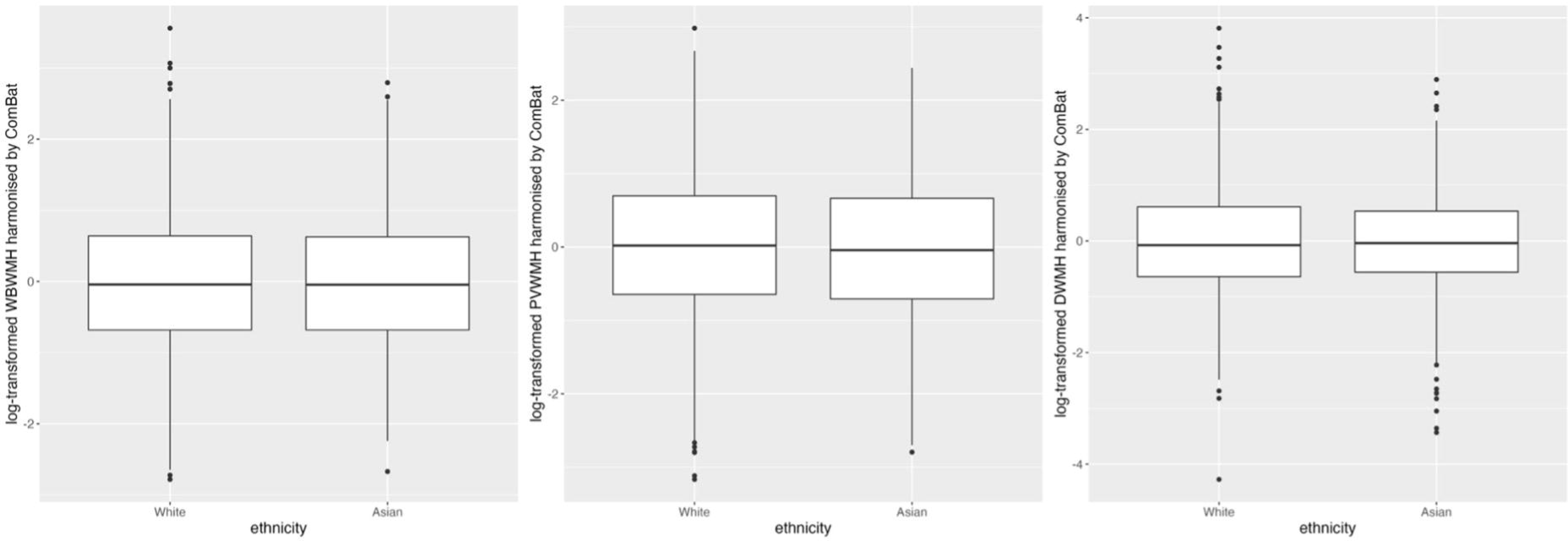
- The differences in WMH volumes between White and Asian participants. WBWMH – whole brain white matter hyperintensities; PVWMH – periventricular white matter hyperintensities; DWMH – deep white matter hyperintensities.

Interaction terms between ethnicity and risk factors with a p-value < 0.1 were found for hypertension (WBWMH), and for stroke and hypercholesterolemia (WBWMH/PVWMH/DWMH). We further investigated these associations by splitting the whole sample into White and Asian subsamples (Table 4). The association between hypertension and higher WBWMH volumes was statistically significant in the White subsample only (β = 0.074, p = 0.001). The associations between hypercholesterolemia and less WBWMH (β = −0.081, p=0.021) and PVWMH (β = −0.084, p=0.039) volumes were significant in the Asian subsample only. The associations of stroke with higher WBWMH (β = 0.124, p = 0.011) and DWMH (β = 0.176, p = 0.026) volumes were statistically significant in the White subsample only.

**Table 4.** Differences in the associations of risk factors with WMH between Whites and Asians.

| Risk Factors | WMH measure | Whites |  |  | Asians |  |  |
| --- | --- | --- | --- | --- | --- | --- | --- |
| | | $\beta$ | CI lower | CI upper | $\beta$ | CI lower | CI upper |
| Hypertension | WBWMH | 0.074** | 0.031 | 0.116 | 0.016 | -0.084 | 0.115 |
| Stroke | WBWMH | 0.124** | 0.029 | 0.220 | 0.006 | -0.127 | 0.139 |
| Stroke | PVWMH | 0.083 | -0.002 | 0.176 | 0.001 | -0.127 | 0.129 |
| Stroke | DMWH | 0.176* | 0.021 | 0.331 | 0.050 | -0.137 | 0.230 |
| Hypercholesterolemia | WBWMH | -0.005 | -0.044 | 0.034 | -0.081* | -0.164 | -0.002 |
| Hypercholesterolemia | PVWMH | -0.019 | -0.042 | 0.036 | -0.084* | -0.168 | -0.001 |
| Hypercholesterolemia | DWMH | -0.003 | -0.076 | 0.038 | -0.061 | -0.177 | 0.055 |

The table shows regression coefficients for risk factors in White and Asian subsamples. Only factors for which an interaction term between risk factor and ethnicity had $p < 0.1$ are included. \* - $p < 0.05$ , \*\* - $p < 0.01$ . CI - 95% confidence interval. WBWMH – whole brain white matter hyperintensity; PVWMH –periventricular white matter hyperintensity, DWMH –deep white matter hyperintensity.

## Discussion

WMH volumes were examined in relation to vascular risk factors and cognitive function, using samples from five international studies from the COSMIC consortium. Diabetes, hypertension, history of stroke, current smoking, high BMI, physical inactivity and higher alcohol consumption were associated with larger WBWMH volumes. WMH volume, especially in the deep white matter regions, was negatively associated with executive function, and higher DWMH volume was significantly associated with poorer performance in processing speed and executive function. The associations of hypertension and stroke with WMH volumes were stronger in White participants, whereas a negative association between hypercholesterolemia and WMH was stronger in Asians.

Our finding of associations between WMH volumes and cardiovascular factors is consistent with previous studies for diabetes (8), hypertension (28), and BMI (29). A few population-based studies reported no association between alcohol consumption and WMH volumes (30, 31). However, the current study showed a significant association between higher alcohol intake and higher WMH volumes. A systematic review showed a linear association between higher alcohol consumption and higher prevalence of hypertension and heart failure (32). Cardiovascular dysfunction was strongly associated with WMH (9). Some pathology studies found that ethanol may affect endothelial function (33) and cause vascular oxidative stress (34), contributing to the accumulation of WMH.

Current smoking was a significant risk factor for WMH volumes, but not past smoking, consistent with previous findings (35). Interestingly, there is no sufficient evidence to support a difference between former smokers and non-smokers. Furthermore, a previous study also reported a positive association between the progression of WMH and an increase in the pack-years of smoking.(35). At the same time, there was no association between years since quitting and WMH progression among former smokers. However, only 6% of participants quit smoking within six years. Future research should track more smokers to investigate the impact of the duration after quitting smoking on WMH.

The results of the current study suggest that a moderate level of physical activity was sufficient enough to be associated with WMH volume. This is consistent with a systematic review that found less WMH to be associated with more physical activity, but only in individuals without severe neurological disorders (36). The optimal amount of physical activity associated with reduced levels of WMH remains to be determined.

We found executive function and processing speed to be associated with both WBWMH and DWMH, though with marginal statistical significance. Some previous studies have also found these cognitive domains to be associated with lower WMH volumes, in both general population-based and high vascular risk samples (37). However, other research has reported contradictory findings of no association of WMH volumes with either global cognition (38, 39) or any specific cognitive domain (38). In the current study, each of the participating cohorts used different neuropsychological test batteries, and we were only able to use one test for each cognitive domain that was common across cohorts. Since one neuropsychological test may not necessarily be representative of a specific cognitive domain, this lack of a broader range of cognitive measures may have contributed to the marginal nature of the association seen in our study.

The current study did not find significant differences in WMH volumes between White and Asians. However, it is worth noting that the Asian cohorts included in the current study (Korea and Singapore) are from relatively high-income countries. A recent systematic review found a higher presence of moderate to severe WMH in low-income countries (moderate to severe WMH: 28.4%, mean age > 60) than in middle-income countries (moderate to severe WMH: 19.0%, 61.5% articles mean age > 60, 38.5% mean age 55-60) (40). A systematic review included participants with a mean age of 61 years who reported a higher presence of moderate to severe WMH in community-based cohorts in high-income countries (Rotterdam, Austrian, moderate to severe WMH > 65%) (41). However, one study in the US reported that Black had higher WBWMH volumes than White (42). Future research is needed to investigate 1) ethnicity differences in WMH volumes in countries with similar income levels, and 2) The association between WMH volumes and income level using a similar data acquisition protocol.

In the current study, hypertension was more strongly associated with WMH in White participants than in Asians. Some prior community-based studies showed that hypertension was associated with higher WBWMH volumes, particularly in older whites (14, 43). In contrast, a recent Korean community-based study (mean age: 73) and a recent Japanese registry database study (ages > 60) reported no association between hypertension and WMH volumes (44, 45). However, another Korean community-based study found a significant association between WMH volumes and hypertension (46). The classification criteria (systolic blood pressure over 130 mm Hg or diastolic blood pressure over 85 mm Hg, no medical history) differed from the study mentioned above and the current study (systolic blood pressure over 140 mm Hg or diastolic blood pressure over 90 mm Hg, with medical history). Another Japanese cross-sectional study found that an association between hypertension and WMH volumes existed in participants with the mean age of 55, but not in the older subsample (mean age > 65) (47).

As for hypercholesterolemia, the current study found no association between hypercholesterolemia and WMH volumes, which is consistent with both Asian (46) and White (48) large-scale studies. Although some community-based studies showed that higher low-density lipoprotein cholesterol was associated with higher WMH burden (49, 50), one cohort (n = 2635) noted a significant association between higher high-density lipoprotein cholesterol and higher prevalence of cerebral microbleeds (51). In the tests for ethnicity differences, hypercholesterolemia was associated with lower WMH volumes in Asian subsamples. A well-presented review (52) proposed that cholesterol-lowering medication (e.g., statin) may moderate the effects of cholesterol on brain imaging measurement (53) and neurocognitive disorders (54). As for the ethnic differences in response to medicines, a previous report showed that Japanese with lower statin doses led to a similar risk reduction of cardiovascular disease than Whites with higher doses (55). A pharmacokinetic study of statins indicated that the overall quantity of statin successfully reaching the systemic circulation in Asians is double compared to Whites (56), which may contribute to brain imaging measurements. Future studies about hypercholesterolemia should consider the effect of statins, especially in elderly research.

There are some limitations in this study. Firstly, harmonizing non-imaging risk factors in multi-cohort studies is challenging. Some studies collected detailed information, such as self-reports, medical history, and clinical data, while others only acquired self-reports. Regarding lifestyle-related risk factors, each study had different protocols for data collection, particularly for physical activity (e.g., only some studies reported the number of hours per week). Secondly, each cognitive domain performance score was based on the results of a single test, which may not comprehensively represent ability in any given domain. Thirdly, the criteria of hypercholesterolemia only considered total cholesterol. Future studies should focus more specifically on high-density or low-density lipoprotein cholesterol.

## Conclusion

This multi-national study confirmed previous findings on the associations of WMH with vascular risk factors and cognition. Hypertension and stroke were found to have strong associations with higher WBWMH volumes in White participants only. In comparison, hypercholesterolemia was found to be associated with lower WMH volumes in Asians only.

## Supporting information

Suppermentary

## Data Availability

Data were provided by the contributing studies to COSMIC on the understanding and proviso that the relevant study leaders be contacted for further use of their data and additional formal data sharing agreements be made. Researchers can apply to use COSMIC data by completing a COSMIC Research Proposal Form available from https://cheba.unsw.edu.au/consortia/cosmic/research-proposals.

## Acknowledgements

The head of COSMIC is Perminder S. Sachdev, and the study coordinator is Darren M. Lipnicki. The research scientific committee leads the scientific agenda of COSMIC and provides ongoing support and governance; it is comprised of member study leaders. The COSMIC research scientific committee and additional principal investigators are listed at https://cheba.unsw.edu.au/consortia/cosmic/scientific-committee.

## Funding

Funding for COSMIC comes from the National Institute on Aging of the National Institutes of Health (Award number: RF1AG057531). Anstey, K.J. was funded by ARC Laureate Fellowship FL190100011. Henry Brodaty was funded by NHMRC Program Grant

The Funding acknowledgement for Wave 2 PATH cohort and MRI studies are NHMRC GNTs 179805, 157125.

### Funding for the H70-study

The study was financed by grants from the Swedish state under the agreement between the Swedish government and the county councils, the ALF-agreement (ALF965812, ALF 716681, ALF 72660), the Swedish Research Council (11267, 2005-8460, 2007-7462, 2012-5041, 2015-02830, 2019-01096), the Swedish Research Council RAM (2013-8717), the Swedish Research Council (NEAR 2017-00639), the Swedish Research Council for Health, Working Life and Wellfare (2012-1138, 2013-1202, 2018-00471, AGECAP 2013-2300, no 2013-2496), Hjärnfonden/Swedish Brain Foundation (FO2014-0207, FO2016-0214, FO2018-0214, FO2019-0163, FO2020-0235), Alzheimerfonden (AF-554461, AF-647651, AF-743701, AF-844671, AF-930868, AF940139, AF-968441, AF-967865), the Alzheimer’s Association Zenith Award (ZEN-01-3151), and Stiftelsen Handlanden Hjalmar Svenssons forskningsfond [HJSV20190229, HJSV2021039, HJSV2022059].

### Funding for SLAS

Agency for Science Technology and Research (A*STAR) Biomedical Research Council (BMRC/08/1/21/19/567) and the National Medical Research Council (NMRC/1108/2007, NMRC/CIRG/1409/2014)

### Funding for KLOSCAD

A grant from the Korean Health Technology R&D Project, Ministry of Health and Welfare, South Korea (grant number HI09C1379 [A092077]).

## Consent Statement

All human subjects involved in this study provided their informed consent.

