## Supplemental Text for "Risk factors and cognitive correlates of white matter hyperintensities in ethnically diverse populations without dementia: the COSMIC consortium"

**(Supplementary Materials)**

**Authorship:** Keshuo Lin, BEng^1^, Wei Wen, PhD^1^, Darren M. Lipnicki, PhD^1^, Louise Mewton, PhD^1^, Rory Chen, MSc^1^, Jing Du, PhD^1^, Dadong Wang, PhD^2^, Ingmar Skoog MD, PhD^3,4,5^, Therese Rydberg Sterner, PhD^3,4,6^, Jenna Najar MD, PhD^3,4,7^, Ki Woong Kim MD, PhD^8,9,10^, Ji Won Han MD, PhD^8,9^, Jun Sung Kim, PhD^8^ , Tze Pin Ng, MD^11,12^, Roger Ho, MD^13^, Denise Qian Ling Chua, MSc^14^, Kaarin J. Anstey, PhD^15,16,17^, Nicolas Cherbuin, PhD^18^, Moyra E. Mortby, PhD^15,16,17^, Henry Brodaty MD, PhD^1^, Nicole Kochan, PhD^1^, Perminder S. Sachdev MD, PhD ^1,19^, Jiyang Jiang PhD^1^*, for the Cohort Studies of Memory in an International Consortium (COSMIC).

**Affiliations:**

1. Centre for Healthy Brain Ageing, School of Clinical Medicine, University of New South Wales, Sydney, Australia
2. CSIRO Informatics and Statistics, Locked Bag 17, North Ryde, NSW 1670, Australia
3. Neuropsychiatric Epidemiology Unit, Department of Psychiatry and Neurochemistry, Institute of Neuroscience and Physiology, Sahlgrenska Academy, at the University of Gothenburg, Sweden
4. Centre for Ageing and Health (AGECAP) at the University of Gothenburg, Sweden
5. Region Västra Götaland, Sahlgrenska University Hospital, Psychiatry, Cognition and Old Age Psychiatry Clinic, Gothenburg, Sweden
6. Aging Research Center, Department of Neurobiology, Care Sciences and Society, Karolinska Institutet and Stockholm University, Stockholm, Sweden
7. Section Genomics of Neurodegenerative Diseases and Aging, Department of Human Genetics, Amsterdam Universitair Medische Centra, Amsterdam, the Netherlands
8. Department of Neuropsychiatry, Seoul National University Bundang Hospital, Seongnam, Korea
9. Department of Psychiatry, Seoul National University College of Medicine, Seoul, Korea
10. Department of Brain and Cognitive Sciences, Seoul National University College of Natural Sciences, Seoul, Korea
11. Khoo Teck Puat Hospital, Singapore
12. Geriatric Education and Research Institute, Ministry of Health, Singapore
13. Institute for Health Innovation and Technology (iHealthtech), National University of Singapore
14. Department of Psychological Medicine, National University of Singapore
15. School of Psychology, University of New South Wales
16. Neuroscience Research Australia
17. Ageing Futures Institute, University of New South Wales
18. National Centre for Epidemiology and Population Health, College of Health and Medicine, Australian National University, Canberra, Australia
19. Neuropsychiatric Institute, The Prince of Wales Hospital, Sydney, Australia.

*** Corresponding author:**

Dr. Jiyang Jiang. Level 1, AGSM building (G27), UNSW Kensington, Sydney NSW 2033,

Australia. E:, T: +61 2 9385 0461.

### Supplementary Text 1: Standardisation of neuropsychological test scores across different studies

Standardisation of neuropsychological test scores across different studies was implemented by the following steps:

1. Calculating the average age, sex (female coded as 0, male coded as 1) and education years based on the data in the five included studies (average age = 72.18 years, average sex = 0.46, average education year = 11.99 years)
2. Within each study, raw MMSE and domain scores were transformed to have a Gaussian (or normal) distribution, with the transformed value having the same percentile value as the original value in the original distribution (implemented by using bestNormalize package in R).
3. Using Winsorize score to remove outliers (+/-3SD)
4. In each cohort, raw MMSE or domain scores were used as the dependent variable. Age, sex and education year were used as the independent variables. Linear regression models were run to estimate the regression coefficients for age, sex and education year in each cohort.
5. In each cohort, apply corresponding regression coefficients calculated in step 4 to average age, sex and education year calculated in step 1 to predict average MMSE or domain scores and associated standard error (SE) for the cohort.
6. Standardise MMSE or domain scores were calculated as

$$\frac{individual raw MMSE or domain scores-predicted average MMSE or domian score (step 5)}{Standard error (step 5)}$$

### Supplementary Text Table 1: Neuropsychological test to represent cognitive domains in each cohort

|  | Memory | Language^a^ | Processing Speed | Executive Function |
| --- | --- | --- | --- | --- |
| H70-study | Memory-in-Reality Test | Animals |  |  |
| KLOSCAD | CERAD 10-word list recall test | Animals | TMTA (360s) | TMTB (360s) |
| MAS | RAVLT trial 7 | Animals | TMTA | TMTB |
| PATH | California Verbal Learning Test (recall of first list) |  | TMTA (300s) | TMTB (300s) |
| SLAS | RAVLT trial 7 | Animals | TMTA | TMTB |

CERAD - Consortium to Establish a Registry for Alzheimer’s Disease neuropsychological assessment battery; RAVLT - Rey Auditory Verbal Learning Test; TMTA – Trail Marking Test A; TMTB – Trail Marking Test B. ^a^ Semantic fluency test.

### Supplementary Text 2: Comparisons between harmonisation methods

As with other multi-site imaging studies, we faced the challenge of MRI scanner differences. Recent studies have attempted to harmonise T1- and diffusion-weighted imaging data and their derived measures (1-3), and the effect of pre-processing strategies on WMH measures has been investigated (4). Biases in neuroimaging measures may come from various sources, including different scanner manufacturers, magnetic field strength, and parameters (5). Promising results in harmonising various imaging measures have been shown by ComBat and its variants (4).

*Methods* - Age main effect (6) and Kolmogorov-Smirnov tests (7) were used to compare the performance of different harmonisation methods in the current study. Studies have shown that age is a strong predictor for WMH volumes (8-10). Therefore, harmonisation methods that resulted in maximal age effects (higher regression coefficients and lower p-values) on WMH were considered superior in the current study (6). The Kolmogorov-Smirnov test (K-S test) is a nonparametric test for the similarity of two distributions. Harmonisation methods that resulted in higher p-values in K-S tests, demonstrating similar distributions, were regarded as better (7). P-values of K-S tests between each pair of scanners were calculated for each harmonisation method. The minimal p-values among whole brain WMH (WBWMH), PVWMH, and DWMH, were used to generate the heatmap (Supplementary Text Figure 2), using the scale_fill_gradientn function in ggplot2. ComBat (4) and CovBat (11) models were implemented with the R packages neuroComBat and CovBat, respectively. ComBat-GAM (12) was implemented with the Python package neuroHarmonize. WMH volumes were logarithm-transformed before harmonisation, because of the non-Gaussian distribution. Age, age-squared, and sex were used as biological factors to preserve in ComBat-GAM models. Age and sex effects were preserved in CovBat and ComBat models.

*Result* - Of the three harmonisation methods (ComBat, ComBat-GAM, CovBat), WBWMH, PVWMH, and DWMH volumes harmonised with ComBat showed the strongest age effects (β = 0.012 – 0.013, p = 2.6×10^-7^ – 3.1×10^-15^), after controlling for sex. Compared to age effects on raw WBWMH volumes before harmonisation (β = 0.170, p < 2.2×10^-16^), age effects on harmonised WBWMH volumes were not as strong (β = 0.011– 0.013, p = 3.1×10^-15^– 1.2×10^-13^), but were smoother and more linear, suggesting a better association (Supplementary Text Figure 1). All three harmonisation methods resulted in higher p-values in K-S tests, compared to raw data without harmonisation (Supplementary Text Figure 2), indicating similar distributions. According to K-S test results, performance of the three harmonisation methods was comparable, but because of the strongest associations with age, further analyses were conducted using WMH measures after ComBat harmonisation.

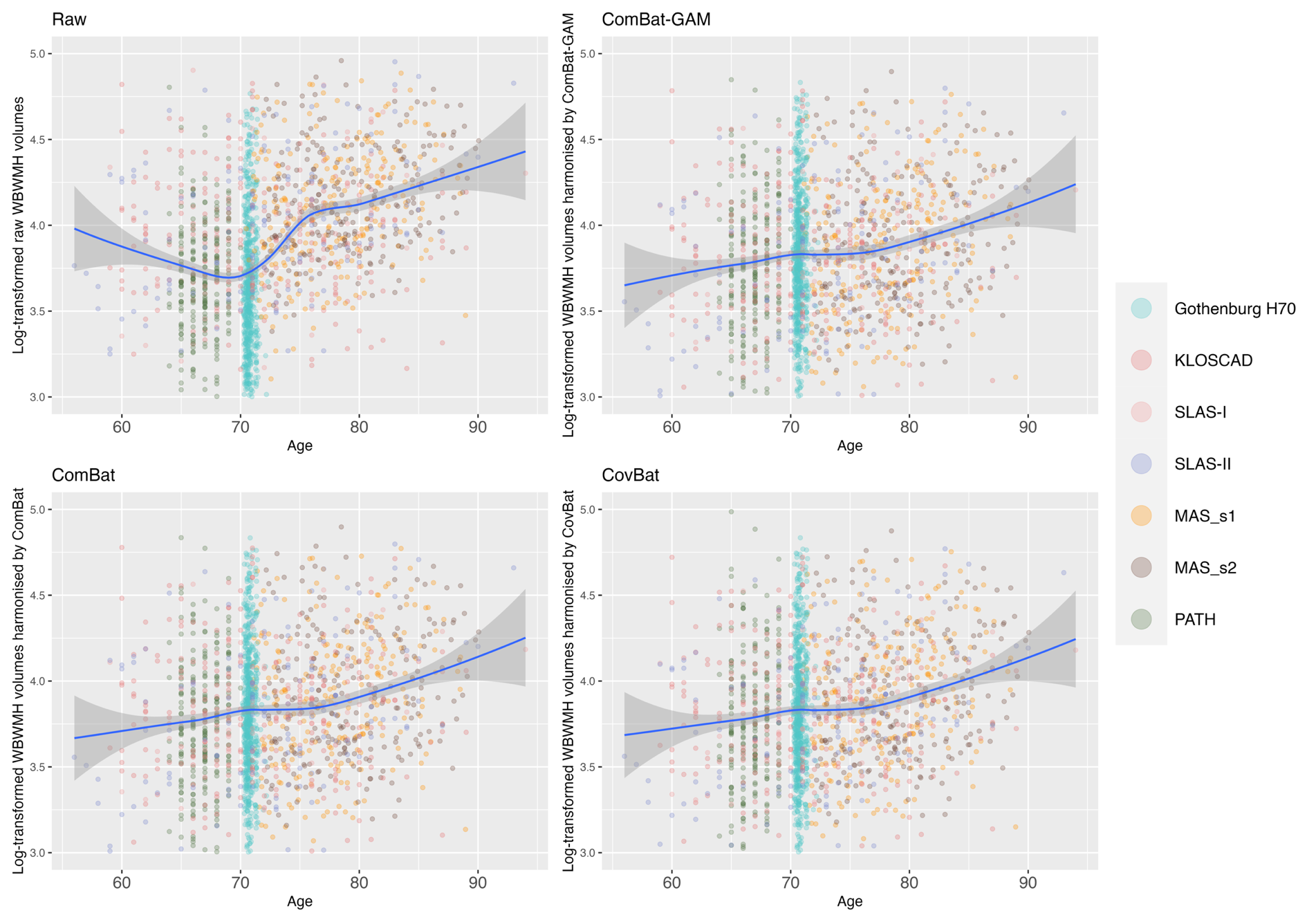

Supplementary Text Figure 1. Age effects on whole brain WMH before and after harmonisation. Locally Estimated Scatterplot Smoothing (LOESS) was applied to generate the curves (span = 0.75). Results showed much smoother age effects after harmonisation, which is more likely to represent the age effects in reality. WBWMH – whole brain WMH; MAS_s1 - MAS Scanner 1; MAS_s2 - MAS Scanner 2.

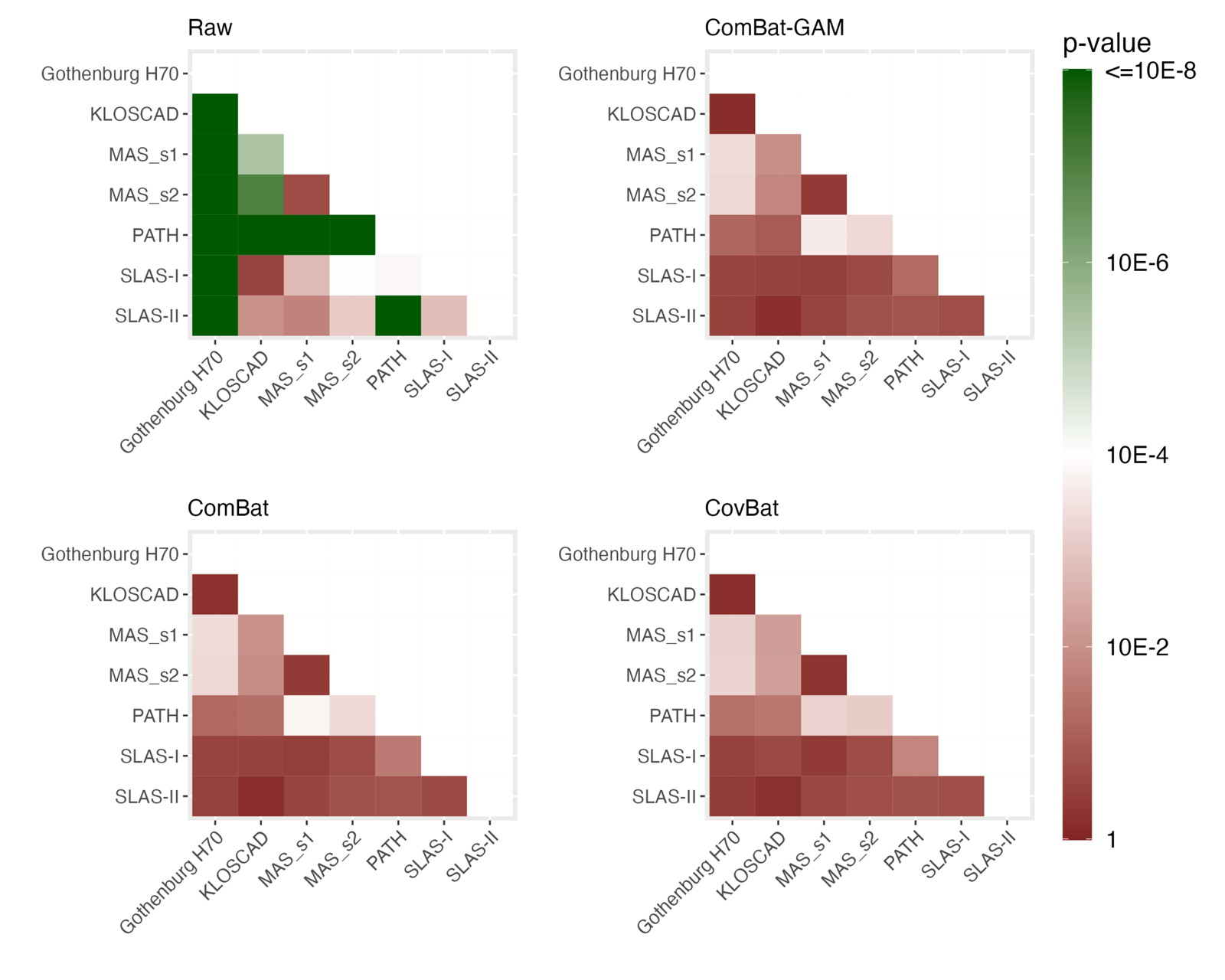

Supplementary Text Figure 2. Kolmogorov-Smirnov (K-S) tests for the distance in distributions of WMH volumes between scanners. P-values of K-S tests between each pair of scanners were calculated for each harmonisation method. Minimum p-values from K-S tests among WBWMH, PVWMH and DWMH volumes were shown. Higher p-values indicate more similar distribution between the pair of scanners, whereas lower p-values indicate more distant distributions. Results showed that all harmonisation methods (ComBat, ComBat-GAM, CovBat) significantly improved the similarity in distributions. MAS_s1, MAS Scanner 1; MAS_s2, MAS Scanner 2.

### Supplementary Text 3: The mediation test procedures

This study also used SEM to explore whether WMH mediates the relationship between vascular risk factors and cognitive function. The following steps were the procedures of the mediation test (mediation test explanation in Supplementary Text Figure 3):

1. examined the total effect between vascular risk factors and cognitive function.

2. evaluated the association between WMH and cognitive function (as risk ~ WMH was explored in the last section)

3. Assessed the mediation effect of WMH on the association between vascular risk factors and cognitive function.

4. Tested the direct effect between vascular risk factors and cognitive function after removing the mediation effect.

All analyses above were conducted controlling for age and sex, using SEM with Lavaan package in R (13). Maximum likelihood estimator with robust standard errors and a scaled test statistic were used. Full Information Maximum Likelihood was used to deal with missing values. Direct and indirect effects were estimated via bootstrapping with 500 samples.

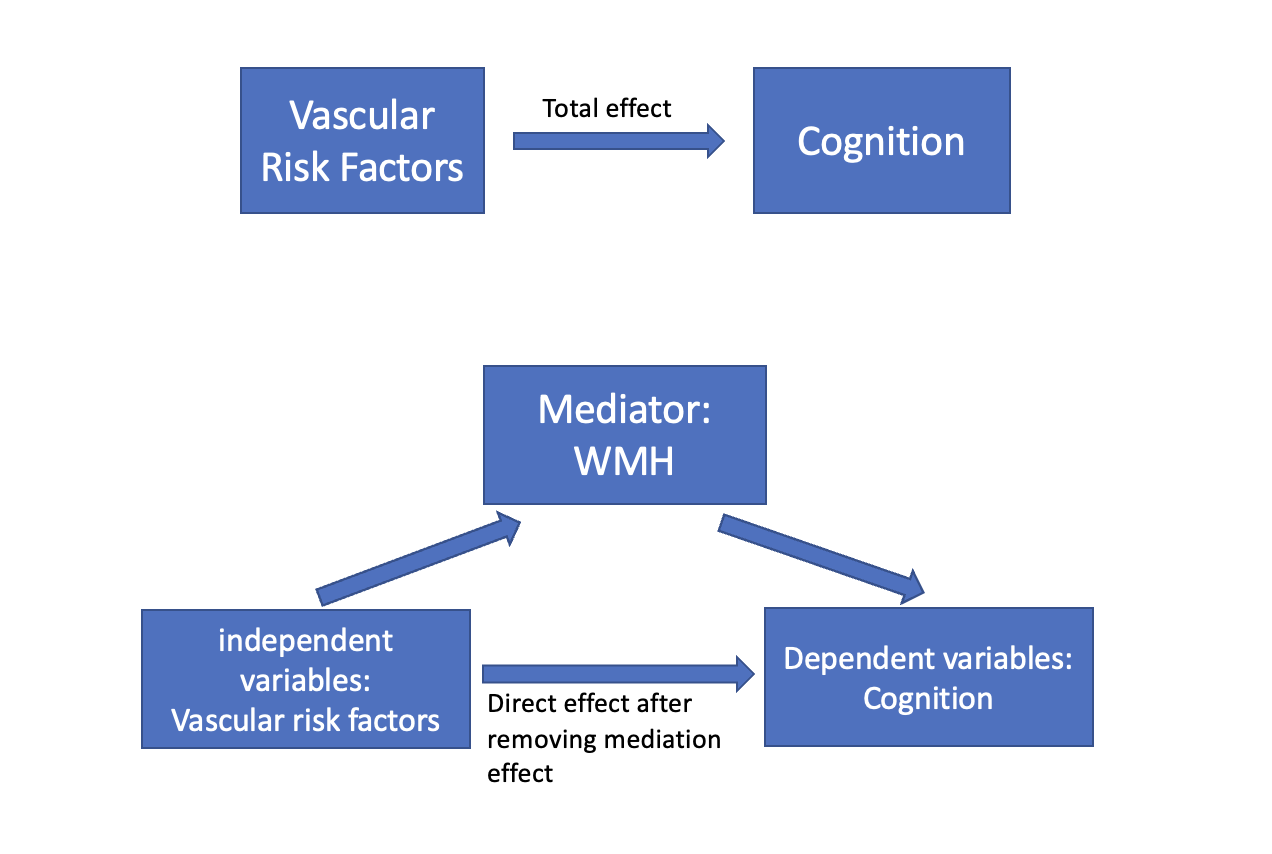

Supplementary Text Figure 3. Mediation test. WMH – white matter hyperintensity.

11.	Andrew A. Chen JCB, Nicholas J. Tustison,, Philip A. Cook RTS, Haochang Shou. Removal of Scanner Effects in Covariance Improves

Multivariate Pattern Analysis in Neuroimaging Data. 2019.

12.	Pomponio R, Erus G, Habes M, et al. Harmonization of large MRI datasets for the analysis of brain imaging patterns throughout the lifespan. Neuroimage 2020;208:116450.

13.	Rosseel Y. lavaan: An R Package for Structural Equation Modeling. J Stat Softw 2012;48(2):1-36.
