## Supplementary material for "Risk factors and cognitive correlates of white matter hyperintensities in ethnically diverse populations without dementia: the COSMIC consortium": Suppermentary

**(Supplementary Materials)**

**Authorship:** Keshuo Lin, BEng^1^, Wei Wen, PhD^1^, Darren M. Lipnicki, PhD^1^, Louise Mewton, PhD^1^, Rory Chen, MSc^1^, Jing Du, PhD^1^, Dadong Wang, PhD^2^, Ingmar Skoog MD, PhD^3,4,5^, Therese Rydberg Sterner, PhD^3,4,6^, Jenna Najar MD, PhD^3,4,7^, Ki Woong Kim MD, PhD^8,9,10^, Ji Won Han MD, PhD^8,9^, Jun Sung Kim, PhD^8^ , Tze Pin Ng, MD^11,12^, Roger Ho, MD^13^, Denise Qian Ling Chua, MSc^14^, Kaarin J. Anstey, PhD^15,16,17^, Nicolas Cherbuin, PhD^18^, Moyra E. Mortby, PhD^15,16,17^, Henry Brodaty MD, PhD^1^, Nicole Kochan, PhD^1^, Perminder S. Sachdev MD, PhD ^1,19^, Jiyang Jiang PhD^1^*, for the Cohort Studies of Memory in an International Consortium (COSMIC).

**Affiliations:**

1. Centre for Healthy Brain Ageing, School of Clinical Medicine, University of New South Wales, Sydney, Australia
2. CSIRO Informatics and Statistics, Locked Bag 17, North Ryde, NSW 1670, Australia
3. Neuropsychiatric Epidemiology Unit, Department of Psychiatry and Neurochemistry, Institute of Neuroscience and Physiology, Sahlgrenska Academy, at the University of Gothenburg, Sweden
4. Centre for Ageing and Health (AGECAP) at the University of Gothenburg, Sweden
5. Region Västra Götaland, Sahlgrenska University Hospital, Psychiatry, Cognition and Old Age Psychiatry Clinic, Gothenburg, Sweden
6. Aging Research Center, Department of Neurobiology, Care Sciences and Society, Karolinska Institutet and Stockholm University, Stockholm, Sweden
7. Section Genomics of Neurodegenerative Diseases and Aging, Department of Human Genetics, Amsterdam Universitair Medische Centra, Amsterdam, the Netherlands
8. Department of Neuropsychiatry, Seoul National University Bundang Hospital, Seongnam, Korea
9. Department of Psychiatry, Seoul National University College of Medicine, Seoul, Korea
10. Department of Brain and Cognitive Sciences, Seoul National University College of Natural Sciences, Seoul, Korea
11. Khoo Teck Puat Hospital, Singapore
12. Geriatric Education and Research Institute, Ministry of Health, Singapore
13. Institute for Health Innovation and Technology (iHealthtech), National University of Singapore
14. Department of Psychological Medicine, National University of Singapore
15. School of Psychology, University of New South Wales
16. Neuroscience Research Australia
17. Ageing Futures Institute, University of New South Wales
18. National Centre for Epidemiology and Population Health, College of Health and Medicine, Australian National University, Canberra, Australia
19. Neuropsychiatric Institute, The Prince of Wales Hospital, Sydney, Australia.

*** Corresponding author:**

Dr. Jiyang Jiang. Level 1, AGSM building (G27), UNSW Kensington, Sydney NSW 2033,

Australia. E:, T: +61 2 9385 0461.

### Abbreviations

MMSE - the total score of the Mini-Mental State Examination

HCH – Hypercholesterolemia

AF – Atrial fibrillation

CVD – Cardiovascular disease

BMI – body mass index

PA – physical activity

WMH – white matter hyperintensity

WBWMH – whole brain white matter hyperintensity

PVWMH – periventricular white matter hyperintensity

DWMH –deep white matter hyperintensity

### Supplementary Table 1. MRI acquisition parameters

|  | H70-study | | KLOSCAD | | MAS Scanner 1 | | MAS Scanner 2 | | PATH | | SLAS-I | | SLAS-II | |
| --- | --- | --- | --- | --- | --- | --- | --- | --- | --- | --- | --- | --- | --- | --- |
|  | T1 | FLAIR | T1 | FLAIR | T1 | FLAIR | T1 | FLAIR | T1 | FLAIR | T1 | FLAIR | T1 | FLAIR |
| In-plane resolution (mm^2^) | 1.02×1.02 | 1×1 | 0.5×0.5 | 0.47×0.47 | 1×1 | 0.488×0.488 | 1×1 | 0.488×0.488 | 1.016×1.016 | 0.898×0.898 | 1×1 | 0.86×0.86 | 0.86×0.86 | 0.86×0.86 |
| Slice thickness (mm) | 1 | 2 | 1 | 3 | 1 | 3.5 | 1 | 3.5 | 2 | 4 | 1 | 3 | 1 | 3 |
| Acquisition matrix | 250×250 | 250×250 | 240×240 | 256×256 | 256×256 | 512×512 | 256×256 | 512×512 | 256×256 | 256×256 | 256×256 | 256×256 | 256×256 | 256×256 |
| Field of view (mm) | 256×256 | 250×250 | 120×120 | 120×120 | 256×256 | 250×250 | 256×256 | 250×250 | 260×260 | 230×230 | 256×256 | 220×220 | 220×220 | 220×220 |
| Repetition time (ms) | 7.2 | 4800 | 8.1 | 9900 | 6.39 | 10 | 6.39 | 10 | 28.05 | 11000 | 2530 | 9500 | 400 | 9500 |
| Echo time (ms) | 3.2 | 280 | 4.6 | 125 | 2.9 | 110 | 2.9 | 110 | 2.65 | 140 | 1200 | 120 | 1.9 | 120 |
| Inversion time (ms) | - | 1650 | - | 2800 | - | 2800 | - | 2800 | - | 2600 | - | 2800 | - | 2200 |
| Flip angle (°) | 9 | 90 | 8 | 90 | 8 | - | 8 | - | 30 | - | 7 | - | 15 | - |

KLOSCAD - Korean Longitudinal Study on Cognitive Aging and Dementia; PATH - The Personality and Total Health (PATH) Through Life study; SLAS- Singapore Longitudinal Aging Study; MAS - Sydney Memory and Aging Study. FLAIR - T2-weighted-Fluid-Attenuated Inversion Recovery

### Supplementary Table 2. Quality control for WMH segmentations

| Cohort | The number of QC exclusions | The reasons of exclusions |
| --- | --- | --- |
| MAS | 23 | - Scan artefacts - Centre area contains WMH - Dura mater is included in the WMH mask - Inaccurate WMH segmentation |
| H70-study | 4 | - Inaccurate WMH segmentation |
| PATH | 30 | - Unusual brain shape affected registration of brain images - Scan artefacts - midline being identified as WMH - Inaccurate WMH segmentation |
| KLOSCAD | 3 | - Scan artefacts |
| SLAS | 18 | - Inaccurate WMH segmentation - Scan artefacts |

QC – quality control.

### Supplementary Table 3. Harmonisation protocols for vascular risk factors in KLOSCAD study

| **Variables** | **Criteria** |
| --- | --- |
| Diabetes | Meeting any of the following criteria is considered a positive diagnosis of diabetes:   - History, - Self-reported current, - Fasting blood glucose criteria are ≥126mg/dL or >7mmol/L. - Non-fasting blood glucose ≥200mg/dL |
| Hypertension | Meeting any of the following criteria is considered a positive diagnosis of hypertension:   - Seated systolic blood pressure ≥140 mmHg or diastolic blood pressure ≥90 mmHg. - Medication - History |
| Hypercholesterolaemia | Meeting any of the following criteria is considered a positive diagnosis of hypercholesterolaemia:   - History, - Self-reported current - Cholesterol are ≥240mg/dL or >6.2mmol/ - Triglycerides ≥200mg/dL or >2.3mmol/L |
| AF | Meeting any of the following criteria is considered a positive diagnosis of AF:   - History - Current status |
| CVD | Meeting any of the following criteria is considered a positive diagnosis of CVD:   - History of any of myocardial infarction, angina, congestive heart failure, arrhythmia, cardiac operation, or other (also having follow-up current status data or age first diagnosed/began medication) - Self-reported current cardiac disease |
| Stroke | Meeting any of the following criteria is considered a positive diagnosis of Stroke:   - History of stroke (sometimes indicated only by having data for a follow-up current status), cerebral infarction, cerebral haemorrhage, TIA, or cerebral ischaemia. - “Something like stroke”. |
| Alcohol | Amount of alcoholic drinks consumed in standard units per week (continuous). |
| Physical Activity | 1+ day/week performing light exercise (such as stepper in-house, slow social dance, golf on the cart, bowling, walking at a speed of 3 to 5 km/hr, free gymnastics or calisthenics; setting-up exercises; rhythmic gymnastics) = 1, and either moderate (biking at speeds of over 16 km/hr, club-dragging golf, slow swimming, fast walking at a speed of 6 km/hr, doubles tennis, fast ballroom dancing) or vigorous exercise (skating, climbing, running, singles tennis, skiing, intense aerobics) = 2 (regardless of time spent). None of these or only light activities (writing, typing, walking slowly at speeds of less than 3 km/hr) = 0. |
| Smoking | “Are you smoking now?” |

### Supplementary Table 4. Harmonisation protocols for vascular risk factors in SLAS

| Variables | Criteria |
| --- | --- |
| Diabetes | Meeting any of the following criteria is considered a positive diagnosis of diabetes:   - Fasting blood glucose criteria are ≥126mg/dL or >7mmol/L. - Treatment - History |
| Hypertension | Meeting any of the following criteria is considered a positive diagnosis of hypertension:   - Blood pressure criteria are seated systolic blood pressure ≥140 mmHg or diastolic blood pressure ≥90 mmHg. - Medication - History |
| Hypercholesterolaemia | Meeting any of the following criteria is considered a positive diagnosis of hypercholesterolaemia:   - History - Treatment - Triglycerides ≥200mg/dL or >2.3mmol/L |
| CVD | Meeting any of the following criteria is considered a positive diagnosis of AF:   - Heart attack - Congestive heart failure - cardiovascular disease - Atrial fibrillation |
| Stroke | History of stroke or regular medication for stroke |
| Alcohol | Drink = never or rarely, less than 1 per week, more than 1 per week but less than 1 per day, 1-2 per day. 97.1% of participants chose never or rarely (considered as non-drinkers). Therefore, we excluded this cohort. |
| Physical Activity | Response options are “never/hardly ever”, “sometimes”, “often”.  Moderate activity (scrubbing, polishing car, dancing, golf, cycling, decorating, lawn mowing, leisurely swimming) “sometimes” = 1. Vigorous activity (running, hard swimming, tennis, squash, digging, cycle racing) “sometimes” or “often” = 2. Less frequent participation in these, only mild activity (walking, woodwork, gardening, bike repairs, playing pool, general housework), or less = 0. |
| Smoking | “Are you smoking now?” |

### Supplementary Table 5 Harmonisation protocols for vascular risk factors in the H70-study

| Variables | Criteria (meeting any is sufficient) |
| --- | --- |
| Diabetes | Meeting any of the following criteria is considered a positive diagnosis of diabetes:   - Treatment - History |
| Hypertension | Meeting any of the following criteria is considered a positive diagnosis of hypertension:   - Blood pressure criteria are seated systolic blood pressure ≥140 mmHg or diastolic blood pressure ≥90 mmHg. - Medication - History |
| Hypercholesterolaemia | Meeting any of the following criteria is considered a positive diagnosis of hypercholesterolaemia:   - History - Triglycerides ≥200mg/dL or >2.3mmol/L - Cholesterol are ≥240mg/dL or >6.2mmol/L |
| AF | Meeting any of the following criteria is considered a positive diagnosis of AF:   - History - Current status |
| CVD | Meeting any of the following criteria is considered a positive diagnosis of CVD:   - Heart attack - Angina - Atrial fibrillation |
| Stroke | History of stroke or regular medication for stroke |
| Alcohol | Drinks per week calculated from consumption in grams/week using 10 g = 1 drink |
| Physical Activity | “almost nothing” or “mainly sedentary, sometimes a walk or easy gardening etc” = 0; “light physical activity 2-4 hours/week” = 1; “moderate physical activity 1-2 hours/week” or “moderate physical activity at least 3 hours/week” or “high physical activity several times/week” = 2. |
| Smoking | “Are you smoking now?” |

### Supplementary Table 6. Harmonisation protocols for vascular risk factors in PATH study

| Variables | Criteria (meeting any is sufficient) |
| --- | --- |
| Diabetes | Meeting any of the following criteria is considered a positive diagnosis of diabetes:   - Treatment - History |
| Hypertension | Meeting any of the following criteria is considered a positive diagnosis of hypertension:   - Blood pressure criteria are seated systolic blood pressure ≥140 mmHg or diastolic blood pressure ≥90 mmHg. - Medication - History |
| Hypercholesterolaemia | Meeting any of the following criteria is considered a positive diagnosis of hypercholesterolaemia:   - Cholesterol are >6.2mmol/L - Triglycerides are >2.3mmol/L |
| CVD | “Do you have heart trouble?” |
| Stroke | “Have you ever suffered a stroke?” |
| Alcohol | Drinks per week calculated using frequency (never, not in last year, monthly or less = 0; 2-4 per month = 0.75; 2-3 per week = 2.5; 4+ per week = 6.34c) and amount typical for a drinking day (1 or 2 = 1.5; 3 or 4 = 3.5; 5 or 6 = 5.5; 7 to 9 = 8; 10+ = 10). |
| Physical Activity | Response options are “never/hardly ever”, “about 1-3 times a month”, “once or twice a week”, “3 times a week or more”.  Moderate activity (scrubbing, polishing car, dancing, golf, cycling, decorating, lawn mowing, leisurely swimming) “once or twice a week” or “3 times a week or more” = 1. Vigorous activity (running, hard swimming, tennis, squash, digging, cycle racing) “once or twice a week” “3 times a week or more” = 2. Less frequent participation in these, only mild activity (walking, woodwork, gardening, bike repairs, playing pool, general housework), or less = 0. |

### Supplementary Table 7. Harmonisation protocols for vascular risk factors in MAS study

| Variables | Criteria (meeting any is sufficient) |
| --- | --- |
| Diabetes | Meeting any of the following criteria is considered a positive diagnosis of diabetes:   - Fasting blood glucose criteria are ≥126mg/dL or >7mmol/L. - Treatment - History |
| Hypertension | Meeting any of the following criteria is considered a positive diagnosis of hypertension:   - Blood pressure criteria are seated systolic blood pressure ≥140 mmHg or diastolic blood pressure ≥90 mmHg. - Medication - History |
| Hypercholesterolaemia | Meeting any of the following criteria is considered a positive diagnosis of hypercholesterolaemia:   - History - Treatment - Cholesterol are ≥240mg/dL or >6.2mmol/L - Triglycerides ≥200mg/dL or >2.3mmol/L |
| AF | Ever diagnosed |
| CVD | Meeting any of the following criteria is considered a positive diagnosis of CVD:   - Heart attack - Angina - Cardiomyopathy - Valve disease - Arrhythmia - Atrial fibrillation |
| Stroke | Diagnosis of stroke or TIA |
| Alcohol | How often have you drink alcohol in the last year (used for frequency analysis; 1= Not in the last year; 2 = monthly or less; 3 = 2-4 times a month; 4 = 2-3 times a week; 5 = 4-6 times a week; 6 = daily; 777=N/A).  How many standard drinks do you have on a typical day (1=1; 2=2 or 3; 3=4 or 5; 4=6 or 7; 5=8 or more; 777=N/A). |
| Physical Activity | Responses given as times (but not always clear what is intended). Any time indicated for separate activities bowls, golf, dancing, walking, other (Pilates, yoga, tai chi, weights) = 1. Any time indicated for tennis, swimming, jogging, bicycling, aerobics, other relevant = 2. Cases with no time indicated or where the time is clearly less than once a week = 0. Note caveat that swimming may often refer to for leisure rather than for exercise (sometimes noted) and the coding as vigorous may include those for whom swimming is really only moderate or less. |
| Smoking | “Are you smoking now?” |

### Supplementary Table 8. Mediation effects of WMH in the associations between vascular risk factors and cognition

|  | Diabetes | hypertension | HCH | AF | CVD | Stroke | Smoking | BMI | PA | Alcohol |
| --- | --- | --- | --- | --- | --- | --- | --- | --- | --- | --- |
| **WBWMH** | | | | | | | | | | |
| **MMSE** | <0.001 | <0.001 | -0.002 | -0.003 | 0.002 | <0.001 | 0.001 | <0.001 | <0.001 | 0.001 |
| **Memory** | <0.001 | <0.001 | 0.007 | 0.011 | -0.009 | <0.001 | -0.006 | <0.001 | 0.002 | -0.005 |
| **Language** | <0.001 | <0.001 | 0.006 | 0.009 | -0.008 | <0.001 | -0.005 | <0.001 | 0.002 | -0.004 |
| **Processing Speed** | <0.001 | <0.001 | 0.005 | 0.007 | -0.006 | <0.001 | -0.004 | <0.001 | 0.001 | -0.003 |
| **Executive Function** | <0.001 | <0.001 | 0.011 | 0.017 | -0.014 | <0.001 | -0.009 | <0.001 | 0.003 | -0.008 |
| **PVWMH** | | | | | | | | | | |
| **MMSE** | 0.002 | <0.001 | <0.001 | <0.001 | <0.001 | -0.001 | 0.002 | <0.001 | <0.001 | <0.001 |
| **Memory** | -0.005 | -0.001 | 0.002 | <0.001 | <0.001 | 0.003 | -0.007 | <0.001 | 0.002 | -0.001 |
| **Language** | -0.002 | <0.001 | <0.001 | <0.001 | <0.001 | 0.001 | -0.002 | <0.001 | <0.001 | <0.001 |
| **Processing Speed** | 0.002 | <0.001 | <0.001 | <0.001 | <0.001 | -0.001 | 0.002 | <0.001 | <0.001 | <0.001 |
| **Executive Function** | -0.007 | -0.002 | 0.002 | 0.001 | <0.001 | 0.004 | -0.008 | <0.001 | 0.003 | -0.001 |
| **DWMH** | | | | | | | | | | |
| **MMSE** | -0.008 | -0.006 | 0.003 | 0.003 | <0.001 | -0.002 | -0.012 | <0.001 | 0.003 | -0.002 |
| **Memory** | -0.003 | -0.002 | <0.001 | 0.001 | <0.001 | <0.001 | -0.004 | <0.001 | 0.001 | <0.001 |
| **Language** | -0.011 | -0.009 | 0.004 | 0.005 | <0.001 | -0.002 | -0.019 | <0.001 | 0.005 | -0.003 |
| **Processing Speed** | -0.012 | -0.010 | 0.005 | 0.006 | <0.001 | -0.003 | -0.020 | <0.001 | 0.006 | -0.003 |
| **Executive Function** | -0.012 | -0.009 | 0.004 | 0.005 | <0.001 | -0.003 | -0.019 | <0.001 | 0.005 | -0.003 |

The table lists all the regression coefficients. P-values for the associations were shown as * - p-value <0.05. No mediation effects of WMH were found in the associations between vascular risk factors and cognition. MMSE - the total score of the Mini-Mental State Examination; WBWMH – whole brain white matter hyperintensity; PVWMH –periventricular white matter hyperintensity, DWMH –deep white matter hyperintensity.
